# Cardiorespiratory Fitness and Neuromuscular Performance in Patients Recovered from COVID-19

**DOI:** 10.1101/2021.01.11.20248930

**Authors:** Murillo Frazão, Amilton da Cruz Santos, Lucas de Assis Pereira Cacau, Paulo Eugênio Silva, Tullio Rocha Petrucci, Mariela Cometki Assis, Rômulo de Almeida Leal, Cláudia Lúcia de Moraes Forjaz, Maria do Socorro Brasileiro-Santos

**Author notes:** Correspondence: Murillo Frazão, Av. Sen. Ruy Carneiro, 412 - Miramar, João Pessoa - PB, Brazil. Zip code 58032-100.

## Abstract

**Objective:** COVID-19 affects cardiorespiratory and muscular systems, causing dysfunctions that may persist after recovery from the acute infection and treatment. The aim of this study was to evaluate cardiorespiratory fitness and neuromuscular performance in these patients.

**Methods:** Patients recovered from mild (n=31) and severe (n=17) COVID-19 were evaluated and compared to healthy subjects (n=15). All volunteers underwent a maximal cardiopulmonary exercise test with simultaneous acquisition of electromyography (EMG). Power output, oxygen uptake (VO_2_), pulse oxygen (O_2_Pulse), cardiovascular efficiency (ΔHR/ΔVO_2_), ventilation (VE), breathing reserve (BR) and ventilatory efficiency (VE/VCO_2_ slope) were analyzed. From EMG, power output for type Ia and IIa activation as well as total neuromuscular efficiency (Δwatts/Δ%RMS) were determined.

**Results:** Patients with severe COVID-19 presented lower VO_2_, O_2_Pulse and VE than mild COVID-19 patients and healthy subjects (p < 0.05 for all comparisons). No differences in ΔHR/ΔVO_2_, BR or VE/VCO_2_ slope were observed among the groups (p > 0.05 for all comparisons). Type IIa and IIb fibers were activated at lower power output in severe than in mild COVID-19 patients and healthy subjects (p < 0.05). Δwatts/Δ%RMS was lower in severe than in mild COVID-19 patients and healthy subjects (p < 0.05).

**Conclusion:** Patients recovered from severe COVID-19 present low cardiorespiratory fitness, activate glycolytic fibers at low power outputs, and show low neuromuscular efficiency; while patients recovered from mild COVID-19 do not present these sequels.

## Introduction

In late December 2019, a previously unidentified coronavirus, currently named severe acute respiratory syndrome coronavirus 2 (SARS-CoV-2), emerged from Wuhan, China, and resulted in a formidable outbreak in many cities(1). Coronaviruses are found in a variety of birds and mammals throughout the world and have a proclivity for emergence. In the past 20 years, three novel human coronaviruses have emerged: SARS-CoV in 2002, Middle East respiratory syndrome (MERS)-CoV in 2012, and the causative agent of coronavirus disease 2019 (COVID-19), SARS-CoV-2(2).

SARS-CoV-2 is a beta coronavirus that is genetically related to but distinct from SARS-CoV. To gain entry into the host cell, the SARS-CoV-2 glycoprotein binds to the cellular receptor angiotensin-converting enzyme 2(2). The viral response phase starts during the first days of infection, and flu-like symptoms are common (mild phase). Some patients progress to an inflammatory response phase. During this stage, patients develop viral pneumonia, and dyspnea/hypoxia might appear. A minority of COVID-19 patients transit into the third and most severe stage of the illness that manifests as systemic hyperinflammation syndrome and multiorgan dysfunction(3).

In the respiratory system, the virus targets cells lining the respiratory epithelium, causing from an asymptomatic infection to severe end-stage lung disease requiring mechanical ventilation. Disease severity is likely to be a combination of direct virus-induced pathology and the host inflammatory response to the infection(2). In the cardiovascular system, there is evidence of myocardial injury with some patients presenting abnormalities similar to myocarditis(4). In the skeletal muscle, some patients showed malaise, muscle soreness, and elevated levels of blood creatine kinase, which is considered an indicator of muscle damage and inflammatory response (5).

As a result of this multisystemic effect, it is reasonable to suggest that COVID-19 can decrease cardiorespiratory fitness and muscular performance, affecting these functions in a direct association with the disease severity. Additionally, these compromises may persist after the patients have recovered from the COVID-19. However, to date, very few data exist about COVID-19 impact on these capacities after recovery and thus the performance sequels of this illness are still poorly understood. Therefore, the aim of this study was to evaluate cardiorespiratory fitness and neuromuscular performance in patients recovered from mild to severe COVID-19. These data could provide physiological guidance for rehabilitation training programs after COVID-19. We hypothesized that recovery patients present impaired cardiorespiratory and neuromuscular performances in accordance to the disease severity.

## Methods

### Study design

An observational study was carried out. In a single-day evaluation, the patients underwent a cardiopulmonary exercise test (CPET) with simultaneous assessment of muscle electromyography (EMG). This study was approved by the local research ethics committee (Instituto de Educação Superior da Paraíba - IESP, opinion No. 4.132.248, CAAE No. 34234720.0.0000.5184) and was registered in the Brazilian Clinical Trial Registration Platform (Number: RBR-6xqcr4).

### Casuistic

A group of COVID-19 patients who were referred for functional evaluation by CPET at the Exercise Physiology Laboratory of the Federal University of Paraíba from July 4^th^ to 14^th^ were considered eligible for this study. Additionally, a group of healthy subjects paired by age, sex and anthropometric characteristics with the COVID patients was enrolled in the study as a control group.

COVID-19 diagnosis was established by clinical symptoms (fever, fatigue, muscle soreness, cough, dyspnea, etc.) associated with a positive laboratory test (nasal swab or serology) and/or chest tomography (ground-glass opacity). Patients were classified as mild (major clinical symptoms without dyspnea or respiratory failure) or severe (major clinical symptoms with dyspnea or respiratory failure), as postulated by Tian et al(6). Patients who met the following inclusion criteria were enrolled: recovered (less than 30 days) from mild to severe COVID-19. Exclusion criteria were based on comorbidity confounding factors. Thus, patients with critical COVID-19 (i.e. who had required intubation and mechanical ventilation) and those with previous cardiac, pulmonary, neurological, hematological or muscular diseases were excluded.

### Cardiopulmonary exercise test

The technical procedures for CPET followed the American Thoracic Society/American College of Chest Physicians guidelines for cycle ergometer testing(7). The CPET was performed on a CG-04 cycle ergometer (INBRAMED, Porto Alegre, Brazil). Each subject performed a ramp-up protocol, starting with warm-up unloaded pedaling for 2 minutes followed by a workload increment individually selected to achieve maximum effort within 8 to 12 min. Subjects were strongly encouraged by verbal stimuli to achieve maximum effort. The VO2000 (MedGraphics, St. Paul, Minnesota, USA) was used for gas analysis, and it was calibrated according to the manufacturer’s instructions. Data were filtered (mean of 7 points) to avoid noise and analyzed by 10s-averages. Before CPET, a resting spirometry was conducted, in which forced expiratory volume in one second (FEV_1_) was measured (ASMA-1, Vitalograph, United Kingdom) to calculate maximum voluntary ventilation (MVV = FEV_1_ x 35).

For analyses, the following variables were considered: power output, peak oxygen uptake (VO_2_), percentage of predicted VO_2_(8), respiratory exchange ratio at maximal effort (RER), oxygen pulse at maximal effort (O_2_Pulse), cardiovascular efficiency (ΔHR/ΔVO_2_), peak ventilation (VE), breathing reserve used during maxima effort (BR = VE/MVV) and ventilatory efficiency (VE/VCO_2_ slope).

### Electromyography activity

During CPET, neuromuscular activity was analyzed by EMG using a signal acquisition module with a 12-bit resolution A/D converter (EMG800C, EMG System, São José dos Campos, Brazil). Sampling frequency was adjusted to 1000 Hz, frequency band to 20-500 Hz and gain to 1000 times. Bipolar Ag/AgCl self-adhesive surface electrodes were used and placed 20 mm apart (center to center) at the right vastus lateralis (2/3 of the way from the anterior superior iliac spine to the lateral side of the patella), according to Surface Electromyography for the Non-Invasive Assessment of Muscles recommendations(9).

Root mean square (RMS) values were used for analysis. EMG breakpoints were analyzed during the ramp-up protocol, as previously described by Lucía et al(10). The increased EMG amplitude reflects the recruitment of additional motor units (11). Based on this, the first EMG breakpoint was assumed to be type IIa fiber activation, and the second EMG breakpoint was assumed to be type IIb fiber activation (Henneman’s principle)(12). The power outputs at the first and second EMG breakpoints were analyzed (figure 1A). For neuromuscular efficiency analysis, EMG data were normalized by the RMS obtained at the maximum effort (%RMS). Neuromuscular efficiency was determined by the relationship between the power output and EMG (watts/%RMS) at each exercise intensity (25%, 50%, 75% and 100% of maximum power output) (figure 1B), while total neuromuscular efficiency (Δwatts/Δ%RMS) was determined by the relationship between the variation in power output and EMG from unloaded pedaling to maximum exercise intensity.

**Figure 1:**
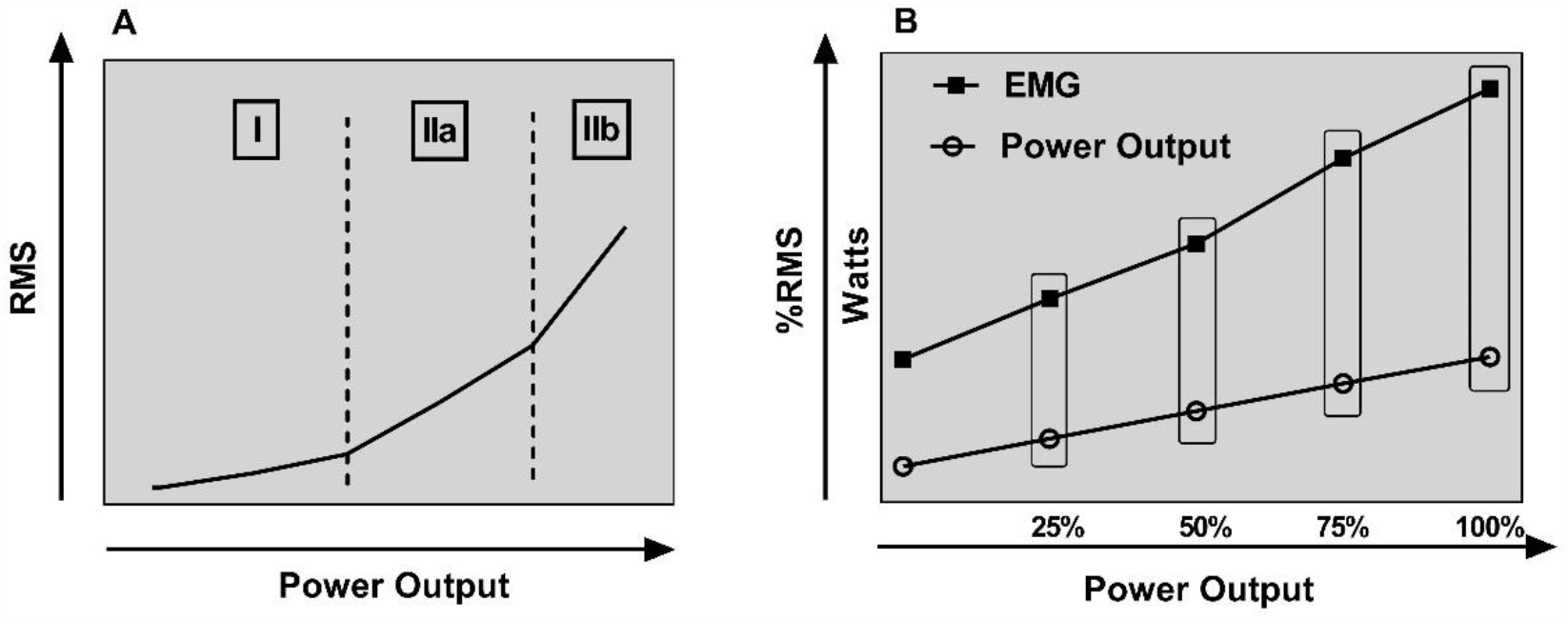
Ilustration of neuromuscular analysis. A: EMG breakpoints. I: activation of type I fibers, IIa: activation of type IIa fibers, IIb: activation of type IIb fibers. B: Neuromuscular efficiency determined by relationship between power output and electromyography (EMG). RMS: root mean square.

### Statistical analysis

Data normality was verified using the Shapiro-Wilk test. Ordinary one-way ANOVA with Tukey’s multiple comparison test were used to evaluate intergroup differences for data with a Gaussian distribution. The Kruskal-Wallis test with Dunn’s multiple comparisons test were used to evaluate intergroup differences for data without a Gaussian distribution. The categorical variables were analyzed by Fisher’s exact test. The effect size was calculated by the F test family (ANOVA: fixed effects, omnibus, one-way) and post hoc type of analysis. The input parameters were as follows: the effect size, f; total sample size; number of groups = 3 and error probability α = 0.05. The effect size, f, was directly calculated from the partial η^2^. The effect size convention was f ≥ 0.1 (small), f ≥ 0.25 (medium) and f ≥ 0.40 (large)(13). A statistical significance value of p ≤ 0.05 was set for all analyses. GraphPad Prism 7.0 and GPower 3.1.9.7 software were used. According to data normality distribution, data are presented as means ± standard deviations or as medians and interquartile ranges and percentages.

## Results

A total of 66 patients were enrolled in the study, but 18 were excluded due to comorbidities (asthma = 9, heart failure = 3, critical COVID-19 = 3, COPD = 2 and fibromyalgia = 1). From the remained 48 patients, 31 had mild and 17 severe disease. The healthy group was composed by 15 subjects. Characteristics of each group are presented in table 1. Anthropometric characteristics are similar among the groups, while COVID symptoms were more frequent in the severe group that also used zinc more frequently as therapy.

**Table 1.** Anthropometric characteristics, main COVID-19 symptoms and main drug therapy.

| **Variable** | **Healthy**<br>**(n = 15)** | **Mild**<br>**(n = 31)** | **Severe**<br>**(n = 17)** | **P value** |
| --- | --- | --- | --- | --- |
| **Age, years** | 43.1 ± 11.2 | 44.5 ± 10.9 | 42.3 ± 10.5 | 0.8463 |
| **Weight, kg** | 80.2 ± 17.3 | 82.8 ± 16.8 | 73.1 ± 17.0 | 0.1782 |
| **Height, cm** | 171.1 ± 10.4 | 160.0 ± 9.4 | 164.1 ± 10.6 | 0.1190 |
| **Sex, M/F%** | 60/40 | 61/39 | 35/65 | 0.1314 |
| **Symptoms, n(%)** |  |  |  |  |
| Fatigue | - | 16 (52) | 12 (70) | 0.2362 |
| Muscle Soreness | - | 12 (39) | 13 (76) | **0.0168** |
| Cough | - | 15 (48) | 10 (59) | 0.5558 |
| Fever | - | 10 (32) | 11 (65) | **0.0383** |
| Headache | - | 5 (16) | 3 (17) | > 0.9999 |
| Diarrhea | - | 6 (19) | 4 (23) | 0.7262 |
| Nausea | - | 4 (14) | 2 (12) | > 0.9999 |
| Anosmia | - | 14 (46) | 9 (53) | 0.1936 |
| Sore Throat | - | 6 (19) | 4 (23) | 0.7266 |
| Dyspnea | - | 0 (0) | 17 (100) | **< 0.0001** |
| Hospitalization | - | 0 (0) | 4 (23) | **0.0122** |
| **Drug therapy, n(%)** |  |  |  |  |
| Hydroxychloroquine | - | 8 (26) | 8 (47) | 0.2014 |
| Antibiotics | - | 23 (74) | 15 (88) | 0.4587 |
| Ivermectin | - | 14 (45) | 12 (70) | 0.1320 |
| Zinc | - | 8 (26) | 10 (59) | **0.0324** |
| Corticosteroids | - | 16 (52) | 6 (35) | 0.3681 |
| Anticoagulants | - | 4 (13) | 2 (12) | > 0.9999 |

M: male and F: female. Mean ± standard deviation.

### Cardiorespiratory fitness

Severe COVID-19 patients presented lower VO2 than mild COVID-19 patients and healthy subjects (1.26 [1.08 – 1.64] vs 1.88 [1.53 – 2.62] vs 2.00 [1.62 – 2.95] L/min; p = 0.0016 compared to mild patients and p = 0.0005 compared to healthy subjects), without a difference between mild COVID-19 patients and healthy subjects (p > 0.9999) (tables 2 and 3). Predicted VO_2_ values were also lower in severe patients than in mild patients and healthy subjects (69.8 ± 10.9 vs 88.6 ± 15.9 vs 97.5 ± 11.5% predicted; p < 0.0001 compared to mild patients and p < 0.0001 compared to healthy subjects), without a difference between mild COVID-19 patients and healthy subjects (p = 0.1025) (table 2). No differences in RER were observed among groups (severe patients: 1.18 [1.12 – 1.26] vs mild patients: 1.16 [1.05 – 1.20] vs healthy subjects: 1.15 [1.07 – 1.15]; p = 0.0746) (table 2).

**Table 2.** Cardiorespiratory fitness parameters.

| *Variable* | **Healthy**<br>**(n =15)** | **Mild**<br>**(n = 31)** | **Severe**<br>**(n = 17)** | **Interaction**<br>**P value** |
| --- | --- | --- | --- | --- |
| ***VO<sub>2</sub>, L/min*** | 2.00 [1.62 – 2.95] | 1.88 [1.53 – 2.62] | 1.26 [1.08 – 1.64]<sup>*#</sup> | **0.0002** |
| ***VO<sub>2</sub>, %predicted*** | 97.5 ± 11.5 | 88.6 ± 15.9 | 69.8 ± 10.9<sup>*#</sup> | **< 0.0001** |
| ***RER*** | 1.15 [1.07 – 1.15] | 1.16 [1.05 – 1.20] | 1.18 [1.12 – 1.26] | 0.0746 |
| ***Power Output, watts*** | 130 [ 103 – 210] | 129 [ 105 – 169] | 81 [ 72 – 131]<sup>*#</sup> | **0.0042** |
| ***O<sub>2</sub>Pulse, mL/beat*** | 14.0 [10.0 – 18.0] | 12.6 [10.6 – 16.0] | 8.0 [7.0 – 12.0]<sup>*#</sup> | **0.0018** |
| ***ΔHR/ΔVO<sub>2</sub>, beats/L/min*** | 45.1 [36.0 – 55.4] | 42.5 [30.5 – 47.1] | 48.7 [32.7 – 73.7] | 0.1630 |
| ***VE, L/min*** | 70.1 [50.3 – 82.0] | 62.5 [47.5 – 77.7] | 47.2 [42.7 – 55.9]<sup>*#</sup> | **0.0174** |
| ***BR, %*** | 39.0 [31.0 – 50.0] | 43.0 [28.0 – 62.5] | 40.0 [36.0 – 46.5] | 0.8636 |
| ***VE/VCO<sub>2</sub> slope*** | 25.3 ± 3.1 | 25.7 ± 4.2 | 25.3 ± 3.1 | 0.9005 |

VO<sub>2</sub>: oxygen uptake, RER: respiratory exchange ratio, O<sub>2</sub>pulse: oxygen pulse, ΔHR/ΔVO<sub>2</sub>: cardiovascular efficiency, VE: minute ventilation, BR: breathing reserve, VE/VCO<sub>2</sub> slope: ventilatory efficiency. \* p ≤ 0.05 compared to healthy subjects. # p ≤ 0.05 compared to mild patients. Mean ± standard deviation. Median [interquartile range].

**Table 3.** Effect size Evaluation.

| Variable | Effect Size (f) | Power (1-β) |
| --- | --- | --- |
| **VO<sub>2</sub>** | Large (0.59) | 0.99 |
| **Power Output** | Large (0.43) | 0.95 |
| **O<sub>2</sub>Pulse** | Large (0.49) | 0.94 |
| **ΔHR/ΔVO<sub>2</sub>** | Medium (0.30) | 0.54 |
| **VE** | Large (0.41) | 0.83 |
| **BR** | Small (0.12) | 0.13 |
| **VE/VCO<sub>2</sub> slope** | Small (0.10) | 0.09 |
| **Ila activation** | Large (0.40) | 0.80 |
| **Ilb activation** | Large (0.48) | 0.92 |
| **Δwatts/Δ%RMS** | Large (0.45) | 0.88 |

VO<sub>2</sub>: oxygen uptake, O<sub>2</sub>Pulse: oxygen pulse, ΔHR/ΔVO<sub>2</sub>: cardiovascular efficiency, VE: minute ventilation, BR: breathing reserve, VE/VCO<sub>2</sub> slope: ventilatory efficiency, Ila activation: power

Severe COVID-19 patients presented lower Power Output than mild COVID-19 patients and healthy subjects (81 [72 – 131] vs 129 [105 – 169] vs 130 [103 – 210] watts; p = 0.0146 compared to mild patients and p = 0.0082 compared to healthy subjects), without a difference between mild COVID-19 patients and healthy subjects (p > 0.9999) (tables 2 and 3).

O_2_Pulse was lower in severe COVID-19 patients than in mild COVID-19 patients and healthy subjects (8.0 [7.0 – 12.0] vs 12.6 [10.6 – 16.0] vs 14.0 [10.0 – 18.0] mL/beat; p = 0.0054 compared to mild and p = 0.0051 compared to healthy subjects), without a difference between mild COVID-19 patients and healthy subjects (p > 0.9999) (tables 2 and 3). No differences in cardiovascular efficiency were observed among groups (severe patients: 48.7 [32.7 – 73.7] vs mild patients: 42.5 [30.5 – 47.1] vs healthy subjects: 45.1 [36.0 – 55.4] beats/L/min; p = 0.1630) (tables 2 and 3).

Severe COVID-19 patients presented lower VE than mild COVID-19 patients and healthy subjects (47.2 [42.7 – 55.9] vs 62.5 [47.5 – 77.7] vs 70.1 [50.3 – 82.0] L/min; p = 0.0400 compared to mild patients and p = 0.0219 compared to healthy subjects), without a difference between mild COVID-19 patients and healthy subjects (p > 0.9999) (tables 2 and 3). No differences in breathing reserve were observed among groups (severe patients: 40.0 [36.0 – 46.5] vs mild patients: 43.0 [28.0 – 62.5] vs healthy subjects: 39.0 [31.0 – 50.0]%; p = 0.8636) (tables 2 and 3). In addition, no differences in VE/VCO_2_ slope were observed among groups (severe patients: 25.3 ± 3.1 vs mild patients: 25.7 ± 4.2 vs healthy subjects: 25.3 ± 3.1; p = 0.9005) (tables 2 and 3).

### Neuromuscular performance

Type IIa fibers were activated at lower power output in severe COVID-19 patients (49 [44 - 67] vs 80 [59 - 93] vs 68 [53 – 88] watts) than in mild COVID-19 patients and healthy subjects; p = 0.0017 compared to mild COVID-19 patients and p = 0.0463 compared to healthy subjects). Type IIb fibers were also activated at lower power output in severe COVID-19 patients (68 [60 - 110] vs 117 [89 - 155] vs 113 [93 - 172] watts) than in mild COVID-19 patients and healthy subjects (p = 0.0047 compared to mild COVID-19 patients and p = 0.0086 compared to healthy subjects). No difference was observed between mild COVID-19 patients and healthy subjects in the activation of either type of fiber (p > 0.9999) (figure 2 and table 3).

**Figure 2:**
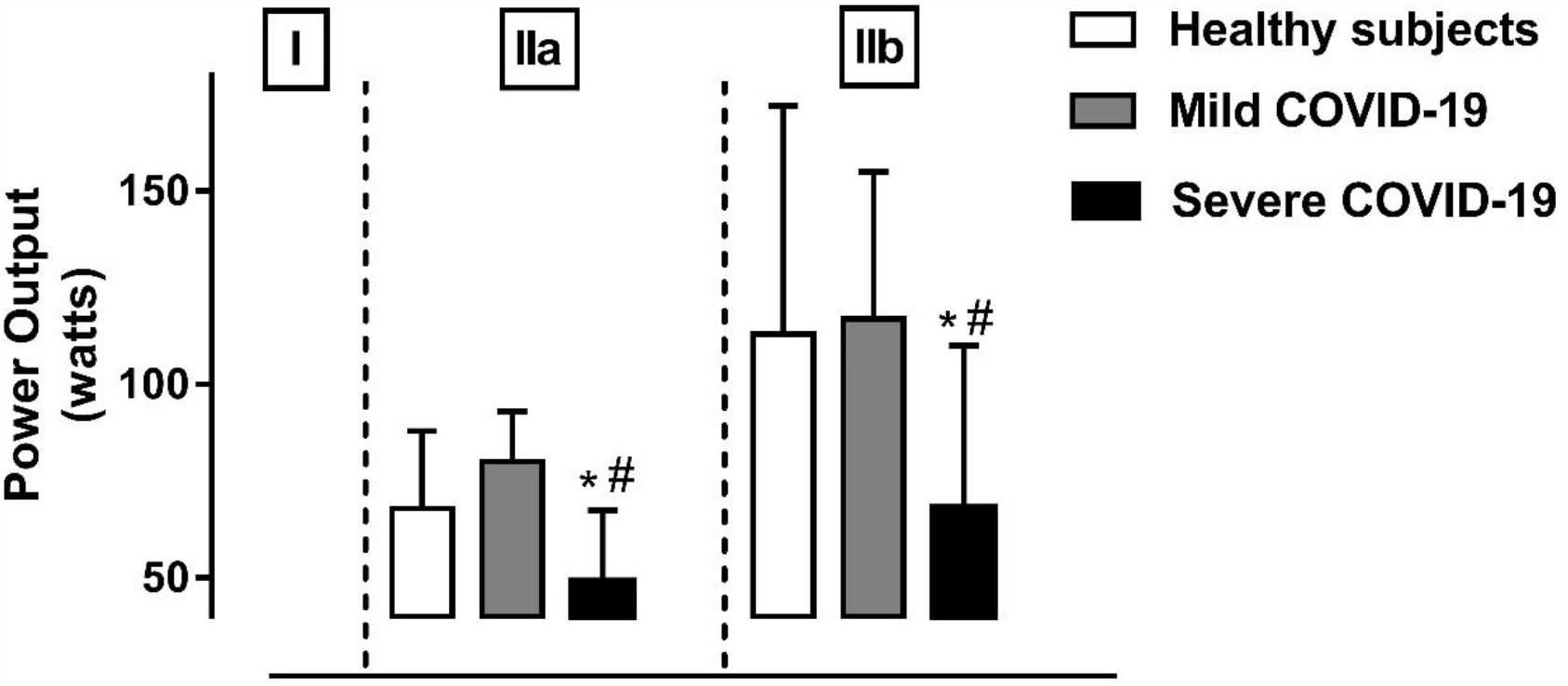
Fiber type activation. I: activation of type I fibers (unloaded pedal), IIa: activation of type IIa fibers, IIb: activation of type IIb fibers. * p ≤ 0.05 compared to healthy. # p ≤ 0.05 compared to mild. Median and interquartile range.

Severe COVID-19 patients presented lower neuromuscular efficiency in all exercise intensities. During light-intensity exercise (25% power output), watts/%RMS = 0.75 [0.49 – 1.03] for severe patients, 1.12 [0.89 – 1.54] for mild patients and 1.44 [1.05 – 2.21] for healthy subjects (p = 0.0350 compared to mild patients and p = 0.0012 compared to healthy subjects, without a difference between mild COVID-19 patients and healthy subjects; p = 0.3145). At moderate-exercise intensity (50% power output), watts/%RMS = 0.76 [0.64 – 1.23] for severe patients, 1.20 [0.89 – 1.47] for mild patients and 1.40 [1.09 – 2.97] for healthy subjects; (p = 0.0484 compared to mild patients and p = 0.0042 compared to healthy subjects, there was no difference between mild COVID-19 patients and healthy subjects; p = 0.4109). During heavy-intensity exercise (75% power output), watts/%RMS = 0.88 [0.71 – 1.14] for severe patients, 1.31 [1.07 – 1.73] for mild patients and1.49 [1.11 – 3.07] for healthy subjects (p = 0.0082 compared to mild patients and p = 0.0016 compared to healthy subjects, there was no difference between mild COVID-19 patients and healthy subjects; p = 0.9489). At maximum exercise intensity (100% power output), watts/%RMS = 0.81 [0.72 – 1.31] for severe patients,1.24 [0.96 – 1.70] for mild patients and 1.30 [1.03 – 2.10] for healthy subjects (p = 0.0359 compared to mild patients and p = 0.0081 compared to healthy subjects, there was no difference between mild COVID-19 patients and healthy subjects; p > 0.9999) (figure 3).

**Figure 3:**
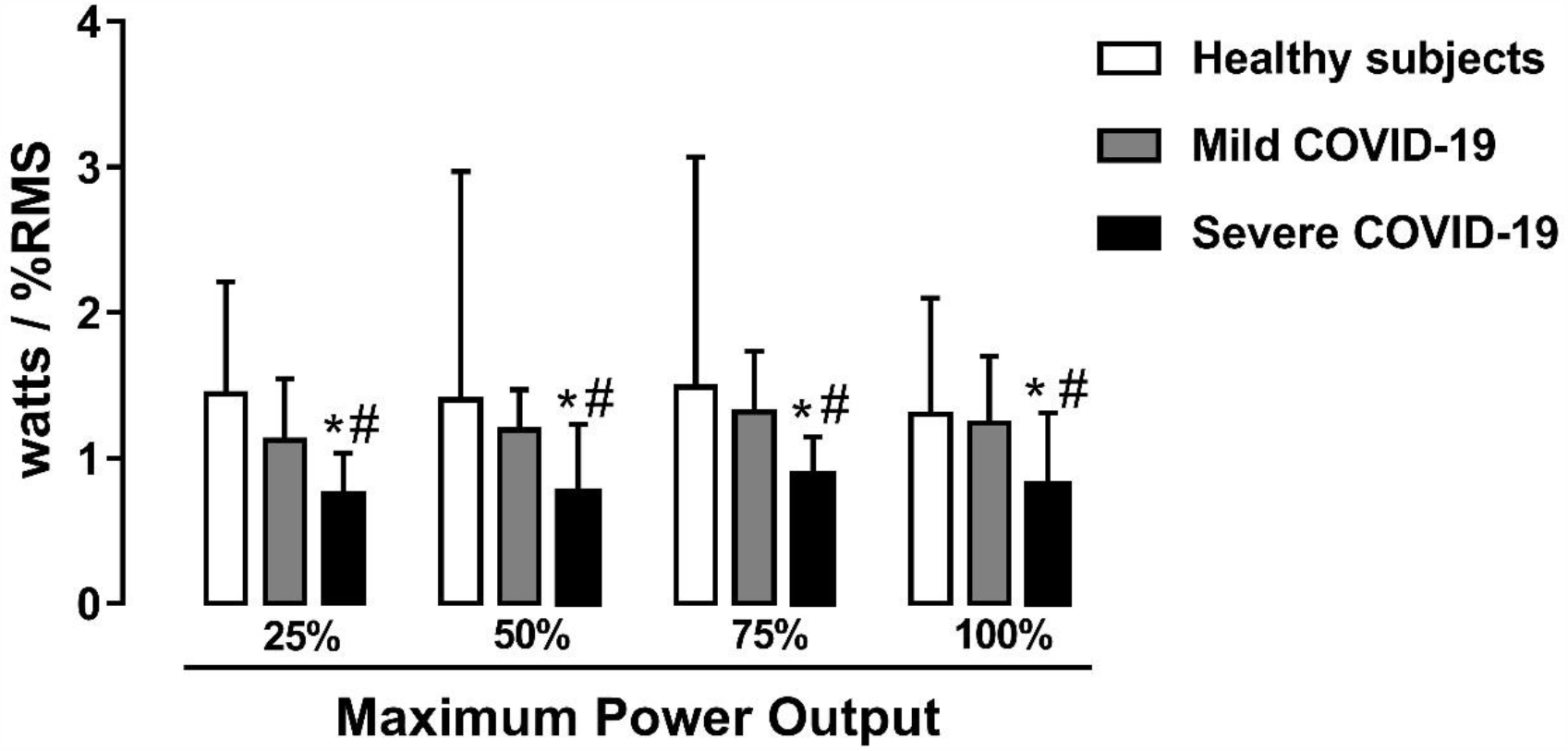
Neuromuscular efficiency in each exercise intensity. * p ≤ 0.05 compared to healthy. # p ≤ 0.05 compared to mild. Median and interquartile range.

Total neuromuscular efficiency was lower in severe COVID-19 patients (0.93 [0.86 – 1.46] vs 1.46 [1.10 – 1.86] vs 1.54 [1.15 – 2.21] Δwatts/Δ%RMS) than in mild COVID-19 patients and healthy subjects (p = 0.0234 compared to mild COVID-19 patients and p = 0.0100 compared to healthy subjects), without a difference between mild COVID-19 patients and healthy subjects (p > 0.9999) (figure 4 and table 3).

**Figure 4:**
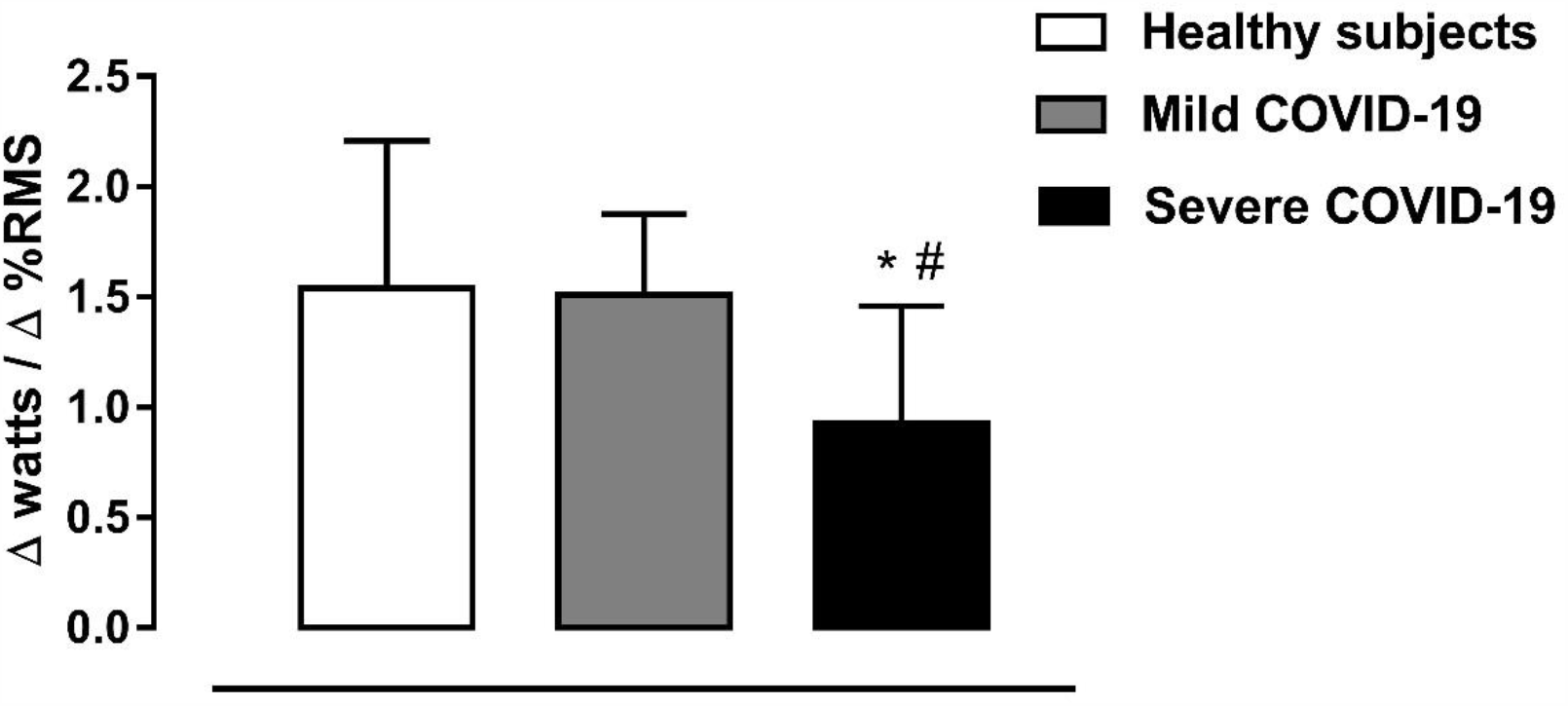
Total neuromuscular efficiency. * p ≤ 0.05 compared to healthy. # p ≤ 0.05 compared to mild. Median and interquartile range.

## Discussion

The main finding of this study was that 1) patients recovered from severe COVID-19 had lower cardiorespiratory fitness, activate muscle fibers Ia and Ib at lower power outputs and present lower neuromuscular efficiency than patients with mild COVID-19 and healthy subjects; while 2) patients recovered from mild COVID-19 had cardiorespiratory fitness and neuromuscular performance similar to healthy subjects.

Peak oxygen uptake reflects functional capacity and has high prognostic value in clinical populations (14)(15). The expected behavior of this variable in patients recovered from COVID-19 is unknown. The present study showed that COVID-19 can have a significant impact on this variable in those patients who had the severe disease but have no effect on those who had only mild symptoms. The hazardous impact observed on VO_2_ may derived from COVID-19’s effects as this disease produces pulmonary function landings (16), myocardial injury (17), and neuromuscular dysfunction (5), but it may also derived from functional impairment imposed by inactivity and hospitalization (18). In a clinical perspective, the reduced VO_2_ observed after severe COVID-19 suggest a worst prognosis for these patients, which should be investigated.

Regarding pulmonary function, it is interesting to note that despite the pulmonary compromise usually seen during COVID-19 evolution, patients with severe symptoms had preserved breath reserve and ventilatory efficiency although peak ventilation was reduced. Xiaoneng et al (19) evaluated resting pulmonary function in 110 patients with COVID-19 discharged from hospital and reported that spirometry values were not affected (forced vital capacity and VEF_1_ > 90% of predicted), while diffusion capacity (64.7% predicted) and total lung capacity (79.2% predicted) were compromised in severe patients. This finding suggests that pulmonary sequalae related to severe COVID may mainly related to lung total volume and gas diffusion, and not to functional volumes that were used during exercise.

Once spirometry values normalize in severe patients and present only a slight reduction in total lung capacity(19), ventilatory performance may have been impaired by respiratory muscle dysfunction. In addition to peripheral muscle, respiratory muscle may present lower performance in severe COVID-19 patients. As demonstrated in healthy(20)(21) and other disease models(22), there is a relationship between peripheral muscle performance and respiratory muscle performance.

Multiple mechanisms of deregulation in pulmonary perfusion exist in COVID-19: the abolition of hypoxic pulmonary vasoconstriction, causing an increase in venous admixture, and excessive pulmonary vasoconstriction and microthrombosis or macrothrombosis, leading to increased deadspace(23). Despite this, the VE/VCO_2_ slope was normal in our sample (even in severe patients). A significant reduction in O_2_pulse was also observed in the group of individuals with severe manifestations; however, the normal ascending behavior of the O_2_pulse curve and preserved ventilatory efficiency reduced the probability of central cardiovascular limitation. One possibility is that those deregulatory mechanisms do not persist after a time of recovery. Another possibility is that only critical patients present significant pulmonary perfusion deregulation.

Thus, individuals recovered from COVID-19 who had severe manifestations had lower functional capacity, without evidence of significant central cardiorespiratory changes, than those recovered from mild COVID-19 or healthy subjects. The reduction in neuromuscular efficiency demonstrated through electromyography suggests that the effort limitation of these individuals is related to a peripheral mechanism.

Severe COVID-19 patients presented lower neuromuscular efficiency, probably due to myositis and a higher inflammatory pattern. Myositis is characterized by the presence of prominent muscle membrane irritability. In myositis, there is invasion and destruction of muscle fibers by cytotoxic T cells(24). SARS-CoV-2 activates inflammatory cytokines, causing inflammatory injury in muscle cells(25). The degree of abnormal muscle membrane irritability is related to disease severity. Mao et al.(26) showed that 11% of the patients had evidence of skeletal muscle injury (creatinine kinase>200 U/L and skeletal muscle pain). Injury was significantly more common in patients with severe disease (19%) than in those with non severe disease (5%). Pizon et al.(5) demonstrated, in a meta-analysis of prevalence based on each neurological manifestation, that muscle injury or myalgia was the most common (19.2%) neurological symptom in individuals with COVID-19.

Type IIa and IIb fibers were activated at much lower power output in severe COVID-19 patients, suggesting worse performance in type I and IIa fibers. Currently, there is insufficient knowledge about muscle histology and fiber-type composition in COVID-19. Independent of any specific pathological process, muscle inactivity causes significant atrophy of all muscle fiber types. Bed rest appears to most strongly induce type I fiber atrophy, accompanied by a fiber-type shift from type I and IIa fibers to type IIb fibers. Taken together, the type of injury, the muscle group affected and the time since injury or rest may all influence how specific muscle fiber types are affected(27).

The pathological process in myopathies results in dysfunction and the dropout of individual muscle fibers located randomly within the motor unit. In myositis, motor neurons and motor axons are not affected. Motor unit action potentials become polyphasic, short in duration and low in amplitude(28). Severe COVID-19 patients presented high EMG activity with low power output because each small motor unit was able to generate only a reduced amount of force, requiring the recruitment of many motor units.

The current study shows a consistent external validity once CPET and EMG are low cost and non-invasive methods, available in the majority of clinical exercise laboratories. However, it is important to point out that we assessed a small sample from a single city in Brazil.

Our study has some limitations. First, a priori sample size calculation was not conducted, however posteriori calculation considering VO_2_ variable resulted in a power of 0.99 and an effect size of 0.54. Sex distribution was different between severe and mild COVID-19 patients, however data presented the same pattern in both sex. Unfortunately, other variables, such as respiratory muscle performance (power and strength) and lactate concentrations that might help to explain the lower capacity observed in severe COVID-19 patients were not assessed in the present study and future studies should evaluate them.

## Conclusions

Patients recovered from severe COVID-19 present low cardiorespiratory fitness, activate glycolytic fibers at low power outputs, and show low neuromuscular efficiency; while patients recovered from mild COVID-19 do not present these sequels.

## Data Availability

We state that all data of this manuscript are available.

## Acknowledgments

The authors thank to the Universidade Federal da Paraíba (UFPB) by grant support to carry out this research, to Exercise Physiology Clinic (CLINAR), João Pessoa - PB, and Infectology and Physiotherapy Clinic (INTERVENT), Aracaju - SE, Brazil.

